# Evaluating the Influence of the Lens Autofluorescence on Adaptive Optics Fluorescence Lifetime Imaging Ophthalmoscopy

**DOI:** 10.64898/2025.12.03.25341556

**Authors:** Ruixue Liu, Xiaolin Wang, Ceren Soylu, Giulia Corradetti, Srinivas R Sadda, Yuhua Zhang

## Abstract

**Purpose:** To assess the impact of lens autofluorescence on adaptive optics fluorescence lifetime imaging ophthalmoscopy (AOFLIO).

**Methods:** Eighteen subjects (n = 18) aged 23 to 68 years with normal chorioretinal health and phakic lenses were imaged using a research-grade AOFLIO instrument. Retinal autofluorescence was excited with a pulsed diode laser (*λ* = 473 nm) and detected in two spectral channels (500–560 nm and 560–720 nm). AOFLIO images were acquired at locations from the foveal center to 10° of the nasal retina. Autofluorescence decay was modeled using bi-exponential and tri-exponential functions, with and without accounting for the early arrival of the lens signal. The contribution of lens autofluorescence was evaluated in relation to age and retinal location.

**Results:** The tri-exponential model that accounted for the early arrival of lens autofluorescence demonstrated superior conformity to the measured autofluorescence decay. The amplitude coefficient of lens contribution was 3 – 4%. However, this component accounted for 19.4 – 29.0% of photon-weighted signals and resulted in a 22.8 – 42.9% overestimation of the mean retinal fluorescence lifetime when uncorrected. Len effect was more pronounced in the short spectral channel. The time shift parameter of lens signal was associated with axial length and age (p = 0.023) and negatively correlated with axial length (p = 0.036).

**Conclusions:** AOFLIO substantially suppresses lens autofluorescence; however, residual lens signal persists and can disproportionately bias retinal autofluorescence lifetime estimates. Accurate correction of the lens component are essential for reliable AOFLIO measurements.

**Translational Relevance:** Minimizing the impact of lens autofluorescence in AOFLIO ensures accurate assessments of metabolic status in the retina and retinal pigment epithelium.

## 1. Introduction

Fluorescence lifetime imaging ophthalmoscopy (FLIO) is an emerging technique for assessing metabolic function of the retina and retinal pigment epithelium (RPE) by measuring the fluorescence lifetimes of endogenous retinal fluorophores.^1-6^ Under one-photon excitation, these fluorophores include lipofuscin, melanolipofuscin, and melanosomes within the RPE, enzymes containing flavin adenine dinucleotide (FAD) or flavin mononucleotide (FMN) located in mitochondria, advanced glycation end products associated with conditions such as diabetic retinopathy, as well as collagen, and elastin in Bruch’s membrane.^3,5,6^ These fluorescent molecules arise as byproducts of cellular metabolism and oxidative stress, or function as coenzymes in energy production, carrying out critical metabolic functions vital to retinal health.^3,5,7^ Alterations in their fluorescence lifetimes have been found to be associated with aging and the development of retinal diseases.^3,4,7^ Thus, accurate measurement of autofluorescence lifetime can potentially detect the functional changes in early stages of retinal pathology. FLIO has been demonstrated to be useful for studying retinal and neurodegenerative diseases, including age-related macular degeneration,^8-11^ diabetic retinopathy,^12^ retinitis pigmentosa,^13^ Stargardt disease,^14,15^ MacTel2,^16^ and Alzheimer’s diseases.^17,18^

Despite its apparent success in clinical applications, the accuracy of FLIO has been found to be substantially impacted by autofluorescence from the human lens, which has a much longer lifetime (on the nanosecond scale) compared to retinal fluorophores (on the order of several hundred picoseconds).^19-21^ The lens’ autofluorescence results in an artificial prolongation of retinal autofluorescence lifetime in FLIO, even with a confocal imaging mechanism.^20,22-24^ To mitigate this artifact, imaging strategies with dual-point confocal scanning and aperture stop separation have been proposed to suppress lens fluorescence,^23,25^ and mathematical models incorporating the contribution of the lens autofluorescence were introduced to improve the fitting accuracy of FLIO.^21,26^

Adaptive optics fluorescence lifetime imaging ophthalmoscopy (AOFLIO) represents a recent advancement that integrates adaptive optics (AO) to improve both lateral and axial resolution.^27-33^ Recently, we have developed an AOFLIO system and demonstrated in vivo assessment of RPE metabolic function at the cellular level.^34-37^ A key feature of AOFLIO is the enhanced confocality afforded by AO correction, which improves rejection of out-of-focus signals. However, the extent to which AO-enhanced confocal imaging suppresses or removes the influence of lens autofluorescence remains uncertain, as does the appropriate method for accurately assessing retinal autofluorescence lifetimes. To address these questions, we evaluated AOFLIO data acquired in human subjects of varying ages with normal chorioretinal health, using different models: a conventional two-term exponential decay and a three-term exponential function with and without accounting for the early arrival time of lens autofluorescence. We further examined whether the early arrival time of the lens correlated with ocular parameters, particularly axial length, raising the possibility of individualized correction strategy. This study aimed to establish a framework for accurate fluorescence lifetime modeling in AOFLIO.

## 2. Methods

The study followed the tenets of the Declaration of Helsinki, complied with the Health Insurance Portability and Accountability Act of 1996, and was approved by the Institutional Review Boards at the University of California at Los Angeles. Written informed consent was obtained from all participants.

### 2.1 Subjects

The participants were recruited from the clinical registry of the Department of Ophthalmology of the University of California, Los Angeles. The inclusion criteria for a normal subject were age greater than 18 years, best-corrected visual acuity (BCVA) of 20/25 or better, refractive error within ±8 D spherical and ±3 D cylindrical, no history of ocular and systemic disease, no significant cataract. A self-report questionnaire was used to ensure participants were in normal chorioretinal health.

The enrolled subjects underwent multimodal ophthalmic imaging, including color fundus photography, fundus autofluorescence imaging (excitation, 488 nm; emission > 600 nm) using an ultra-widefield (UWF™) retinal imager (California, Optos, Inc., Marlborough, MA), and spectral-domain optical coherence tomography (SD-OCT, Spectralis, Heidelberg Engineering Inc., Franklin, MA). Prior to imaging, subjects’ pupils were dilated, and accommodation was paralyzed by instillation of topical tropicamide (1%) and phenylephrine hydrochloride (2.5%) to ensure optimal imaging conditions. A retinal specialist reviewed the images to confirm the retinal health status. Axial length measurements were obtained using an ocular biometer (IOL Master; Carl Zeiss Meditec, Germany).

### 2.2 AOFLIO instrument

The AOFLIO instrument used in this study has been reported elsewhere.^34-37^ For the readers’ convenience, we summarize the technical details here. The AOFLIO was developed based on an adaptive optics scanning laser ophthalmoscope (AOSLO),^38-41^ which operates with a confocal imaging mode using an infrared low coherence light source (Superluminescent diode HP 795, Superlum Ltd, Ireland, λ = 795 nm, Δλ = 15 nm). The AO system consists of a custom designed Shack-Hartmann wavefront sensor and a deformable mirror with 97 actuators (DM97-15, ALPAO SAS, France).

The AOFLIO employs a pulsed diode laser with wavelength at 473 nm (BDS-SM-473, Becker & Hickl GmbH, Berlin, Germany) as the excitation source. The emitted autofluorescence was detected simultaneously in two spectral channels: a short spectral channel (SSC, 500–560 nm) and a long spectral channel (LSC, 560– 720 nm). The arrival times of the autofluorescent photons relative to their excitations were recorded by a time-correlated single photon counting (TCSPC) module (SPC-180nx, Becker & Hickl GmbH, Berlin, Germany) at a frequency of 40 MHz. AOFLIO and AOSLO (λ = 795 nm) were simultaneously acquired through a pupil diameter of 6 mm. The confocal pinholes have a diameter of 150 µm, which corresponds to 5.8 Airy disk diameters (ADD) for the LSC and 7.0 ADD for the SSC, resulting in a lateral resolution of approximately 2 µm and an axial resolution of approximately 70 µm for the AOFLIO. Retinal images were acquired over a field of view of 1.2° × 1.2° and digitized by 512 × 512 pixels at a frame rate of 15 frames per second. For each subject, AOFLIO images were taken at 5 - 7 retinal locations from the foveal center extending to 10 degrees nasally in the horizontal retinal median.

The TCSPC module measured the time intervals between the arrival autofluorescence photons and the closest excitation laser pulses and marked the elapsed time since the recording began. Autofluorescence photons were allocated to a two-dimensional image matrix based on the line- and frame-synchronization signals, and AOSLO pixel clock. To accurately compensate for ocular movements, non-interpolative desinusoidal and substrip-based non-interpolative image registration algorithms^42^ were applied to precisely assign the autofluorescence photons to the corresponding pixel location, ensuring accurate construction of the fluorescence lifetime histogram on each pixel.

### 2.3 Data Analysis

Retinal autofluorescence lifetime data were analyzed using SPCImage software (Version 8.8, Becker & Hickl GmbH, Berlin, Germany). Statistical analyses were carried out using MATLAB (MATLAB R2024a, The MathWorks, Inc., Natick, MA) statistical toolbox.

To evaluate the influence of lens autofluorescence on AOFLIO lifetime measurements, we compared three fluorescence decay models commonly used in conventional FLIO analysis,^26^ including

1. A tri-exponential function accounting for the early arrival of lens autofluorescence photons with a time shift *τ*_*shift*_ (S3-exp):

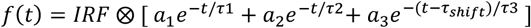
2. A tri-exponential model without accounting for the early arrival of the lens signal (3-exp):

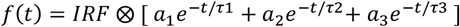
3. A conventional bi-exponential model (2-exp):

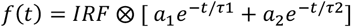

where *IRF* is instrumental response function, *a*_*i*_ denotes the amplitude of the fluorescent component, *τ*_*i*_ is the lifetime of the fluorescent components, and *τ*_*shift*_ is the time shift parameter for the early arrival lens signal.

The mean autofluorescence lifetime was calculated as:

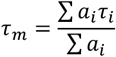

The contribution of each component was assessed using:

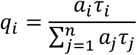

For the tri-exponential models, the mean retinal autofluorescence lifetime was estimated using the first two components (*τ*_1_, *τ*_2_) and their amplitudes (*a*_1_, *a*_2_):^26^

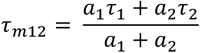

To ensure a reliable estimate of the fluorescence decay, each model was fitted with independently optimized synthetic IRF, whose temporal position and width were refined by nonlinear least-squares optimization jointly with the decay parameters.^43^ Autofluorescence decay was fitted using a moving 13 × 13-pixel binning scheme across the AOFLIO image. Only regions containing more than 2,000 detected photons were included in the analysis. In this study, the photon counts per histogram in the selected regions ranged from 2,000 to 20,616 in the LSC and from 2,000 to 9,363 in the SSC.

For each individual eye, the temporal shift parameter in the S3-exp model was determined by the chi-square (χ^2^) value. The one corresponding to the minimum χ^2^ was considered as the optimal shift.

Statistical analysis was performed to compare fluorescence decay models using one-way analysis of variance (ANOVA), followed by Tukey post hoc tests for pairwise comparisons. Age-related differences and spatial trends were illustrated by stratifying participants into two age groups and plotting autofluorescence lifetime as a function of retinal imaging location. To account for repeated measurements within subjects, linear mixed-effects (LME) models were used to evaluate the effects of age and retinal imaging location on autofluorescence lifetime parameters (τ_3_ and a_3_) derived from the S3-exp model. Additional LME analyses assessed the effects of axial length, retinal imaging location, and age on the time-shift parameter.

In the LME, subjects and measurements were included as random effects. The measurements acquired from the same subject were treated as repeated measurements nested within the subject. Age, axial length, and retinal eccentricity were treated as fixed effects. Model parameters were estimated using restricted maximum likelihood. Regression coefficients (β), representing the estimated change in each outcome per unit change in a predictor while holding other variables constant, were reported with 95% confidence intervals and corresponding P values. Statistical significance was defined as P < 0.05. Model adequacy was evaluated using Akaike and Bayesian information criteria (AIC/BIC), log-likelihood, marginal and conditional R^2^ values, intraclass correlation coefficients (ICC), and variance inflation factors (VIF). Residual diagnostics were performed to assess normality, homoscedasticity, and potential model misspecification.

## 3. Results

**Subject characteristics:** Eighteen subjects (7 males, 11 females; age range, 23–68 years) were enrolled. Imaging was performed at nasal retinal eccentricities from 0° to 10°, yielding a total of 81 retinal locations. Mean refractive error was 0.03 ± 1.38 diopters (range, −7.0 to 1.75 D), and mean axial length was 24.45 ± 1.26 mm. Participants were stratified into two age groups: a younger group (n = 9; mean age, 32.4 ± 4.9 years; range, 23–39 years) and an older group (n = 9; mean age, 59.6 ± 5.1 years; range, 53–68 years).

### 3.1 The S3-exp model showed superior conformity to the autofluorescence decay

The superior fitting accuracy of S3-exp model is demonstrated by the high conformity of the measurement data with the photon decay curve, especially in the rising edge zone. The S3-exp model presented the minimum discrepancy versus the 2-exp and standard 3-exp models (**Fig. 1**).

**Fig. 1.**
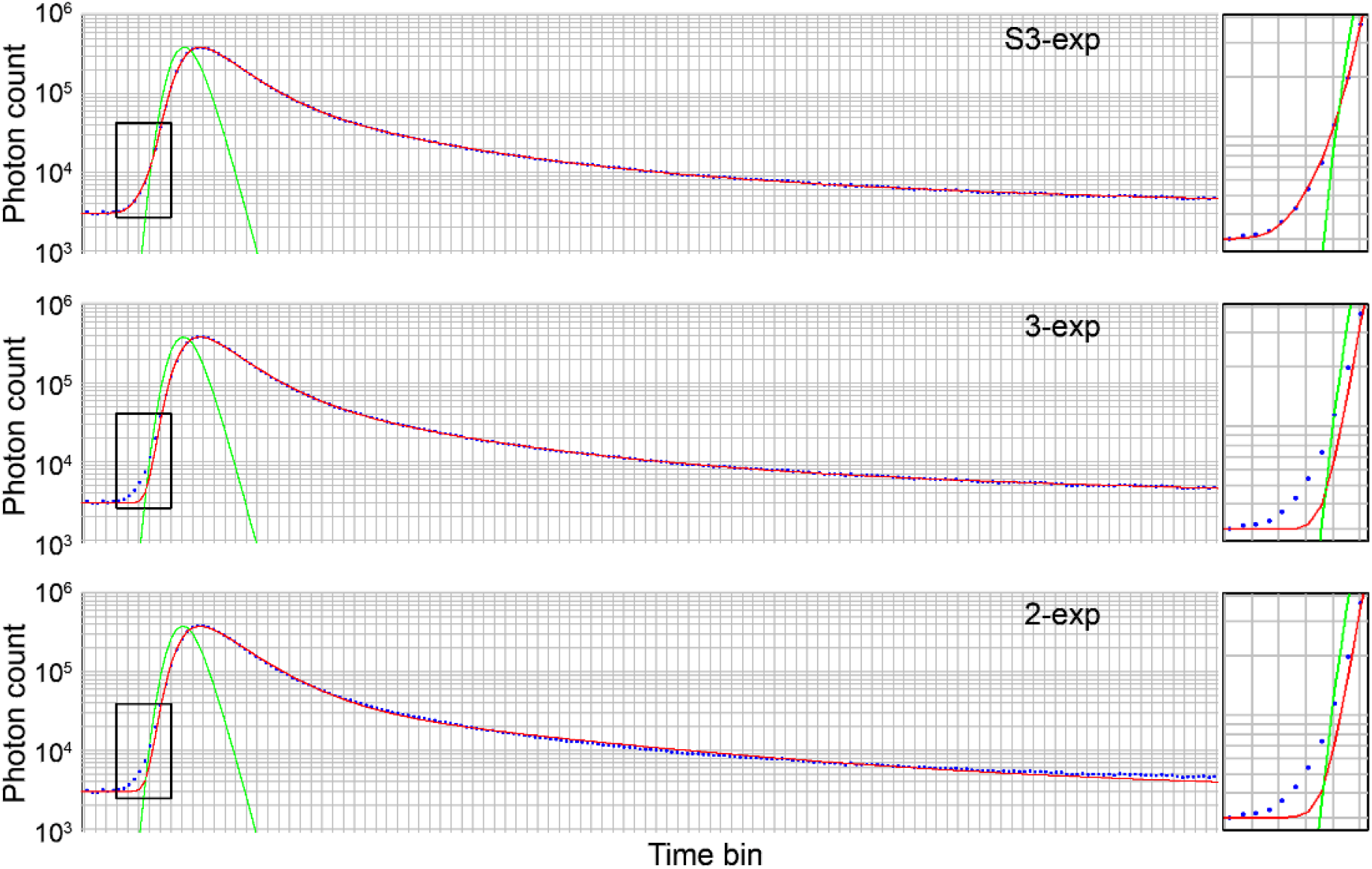
Modeling autofluorescence decay measured by AOFLIO using a bi-exponential function (2-exp), tri-exponential functions with (S3-exp) and without (3-exp) accounting for lens autofluorescence. The boxes show magnified conformity of the AOFLIO measurements with the fitting curve in the rising edge zone. Blue dots are AOFLIO measurements of the photon numbers in the subsequent time bins, representing the time difference between the autofluorescence photons and the excitations. Red lines represent the fitted curves. Green lines indicate the instrument response function.

As shown in **Fig. 2**, in the LSC, the mean χ^2^ was 1.065 for the S3-exp model, compared to higher values of 1.131 and 1.230 for the standard 3-exp and 2-exp models, respectively. In the SSC, the S3-exp model achieved a mean χ^2^ of 1.062, outperforming the standard 3-exp (χ^2^ = 1.135) and 2-exp (χ^2^ = 1.191) models. One-way ANOVA confirmed significant overall differences among these models (p < 0.001 for both channels). Tukey’s post-hoc analysis of pairwise differences revealed the distinct superiority of the S3-exp model.

**Fig. 2.**
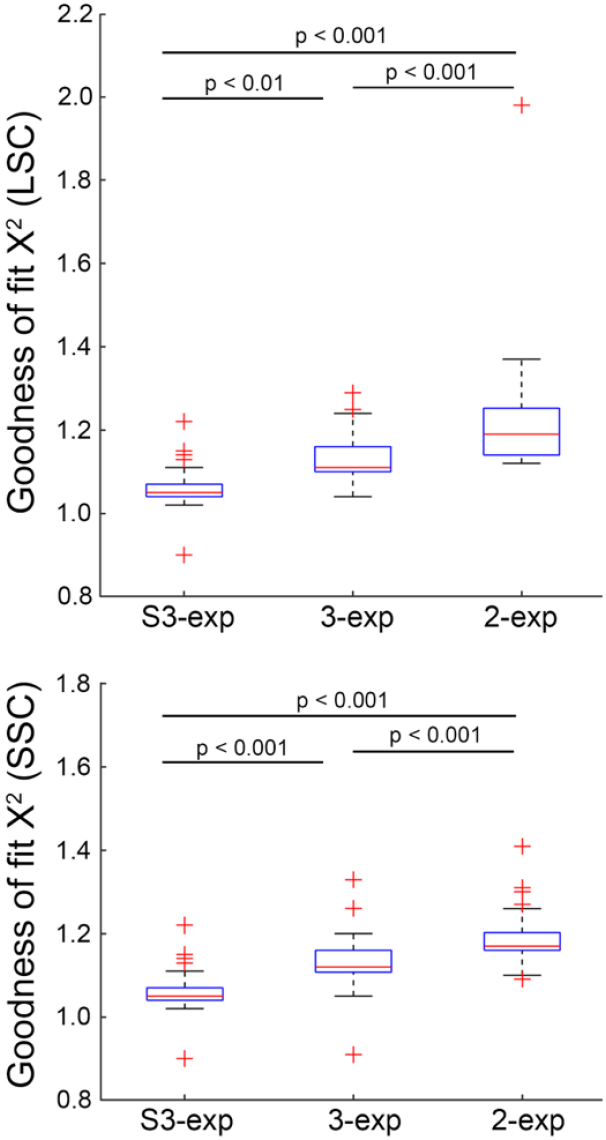
Comparison of the goodness-of-fit (χ^2^) using bi-exponential (2-exp), tri-exponential with (S3-exp) and without (3-exp) accounting for lens signal for estimating the autofluorescence lifetime in the long spectral channel (LSC) and short spectral channel (SSC).

### 3.2 The time shift in the S3-exp model is correlated with the eye’s axial length in the SCC

The mean time shift was 122.0 ± 8.6 ps in the LSC and 132.9 ± 9.8 ps in the SSC. LME modeling disclosed that the time shift was not significantly associated with axial length in the LSC (p = 0.114, **Table 1**), but a positive association in the SSC (p = 0.036, **Table 2**), as shown in **Fig. 3**. It also showed that the time shift was positively associated with age in the SSC (p = 0.023, **Table 2**) but not in the LSC (p = 0.195, **Table 1**). Furthermore, LME demonstrated a consistent dependence of the time shift on retinal imaging location across both channels, with significant decreases toward greater eccentricity (β = −1.70 for both LSC and SSC, all p < 0.001, **Tables 1** and **2**).

**Table 1.** Associations between time shift and axial length and retinal eccentricity (long spectral channel)

| Time shift | $\beta$ | CI- | CI+ | SE | $p$ value | VIF |
| --- | --- | --- | --- | --- | --- | --- |
| Axial length | 1.75 | -0.43 | 3.93 | 1.10 | 0.114 | 1.03 |
| Age | -0.11 | -0.28 | 0.06 | 0.08 | 0.195 | 1.04 |
| Retinal eccentricity | -1.70 | -2.17 | -1.23 | 0.24 | < 0.001 | 1.01 |
| Marginal $R^2$ | 0.349 | | | | | |
| Conditional $R^2$ | 0.605 | | | | | |
| ICC | 0.392 |  |  |  |  |  |

$\beta$ : predictive strength of retinal layer thickness for cone density
CI: 95% confidence interval
SE: standard error
VIF: variance inflation factor
ICC: intraclass correlation coefficients
**These abbreviations apply to Tables 1 through 6.**

**Table 2.** Associations between time shift and axial length and retinal eccentricity (short spectral channel)

| Time shift | $\beta$ | CI- | CI+ | SE | $p$ value | VIF |
| --- | --- | --- | --- | --- | --- | --- |
| Axial length | 2.91 | 0.20 | 5.62 | 1.36 | 0.036 | 1.03 |
| Age | 0.24 | 0.03 | 0.45 | 0.10 | 0.023 | 1.04 |
| Retinal eccentricity | -1.70 | -2.21 | -1.19 | 0.26 | < 0.001 | 1.01 |
| Marginal $R^2$ | 0.388 | | | | | |
| Conditional $R^2$ | 0.628 | | | | | |
| ICC | 0.481 |  |  |  |  |  |

**Fig. 3.**
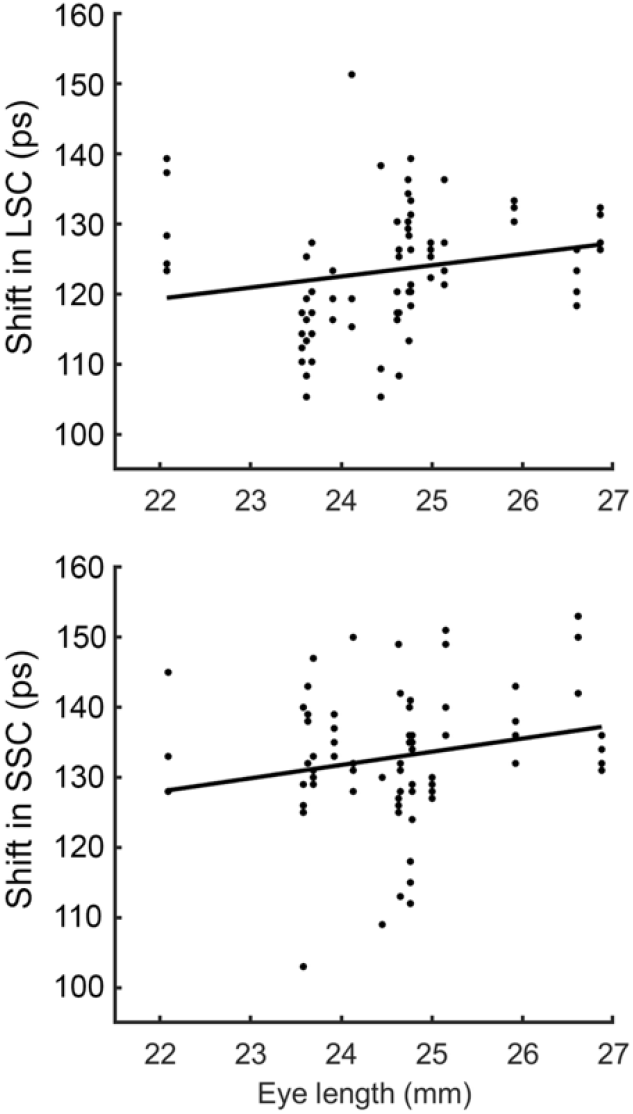
The time shifts in the tri-exponential model accounting for the early arrival of the lens signal versus the axial length of the eye in the long spectral channel (LSC) and short spectral channel (SSC).

### 3.3 Lens autofluorescence increases with age and varies across retinal eccentricity

LME revealed a significant age dependency of the lens fluorescence lifetime (*τ*_3_) of the S3-exp model in both channels (all p ≤ 0.05, **Tables 3** and **4**), also shown in **Fig. 4** top panels. In contrast, the weight *a*_3_ was not influenced by age in either channel (LSC: p = 0.871, SSC: p = 0.989, **Tables 5** and **6, Fig. 4** bottom panels).

**Table 3.** Associations between*τ*_3_ and age and retinal eccentricity (long spectral channel)

| Time shift | $\beta$ | CI- | CI+ | SE | *p* value | VIF |
| --- | --- | --- | --- | --- | --- | --- |
| Age | 7.11 | 0.12 | 14.11 | 3.51 | 0.046 | 1.01 |
| Retinal eccentricity | 9.69 | -3.28 | 22.66 | 6.51 | 0.141 | 1.01 |
| Marginal R<sup>2</sup> | 0.146 |  |  |  |  |  |
| Conditional R<sup>2</sup> | 0.707 |  |  |  |  |  |
| ICC | 0.657 |  |  |  |  |  |

**Table 4.** Associations between*τ*_3_ and age and retinal eccentricity (short spectral channel)

| Time shift | $\beta$ | CI- | CI+ | SE | *p* value | VIF |
| --- | --- | --- | --- | --- | --- | --- |
| Age | 9.86 | 2.16 | 17.57 | 3.87 | 0.012 | 1.01 |
| Retinal eccentricity | -15.39 | -27.54 | -3.24 | 6.10 | 0.014 | 1.01 |
| Marginal R<sup>2</sup> | 0.248 |  |  |  |  |  |
| Conditional R<sup>2</sup> | 0.799 |  |  |  |  |  |
| ICC | 0.733 |  |  |  |  |  |

**Table 5.**
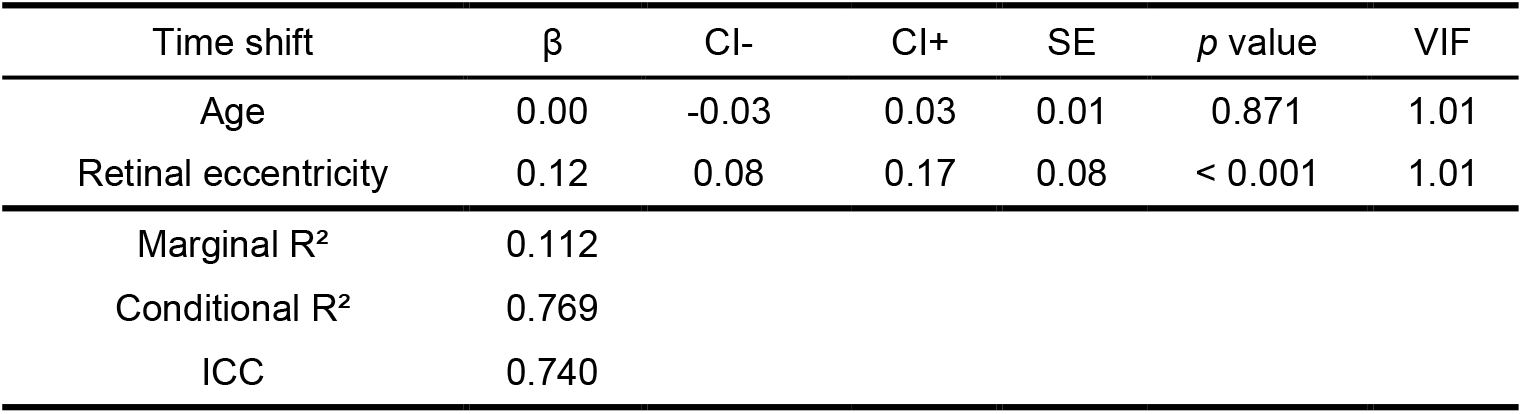
Associations between a3 and age and retinal eccentricity (long spectral channel)

**Fig. 4.**
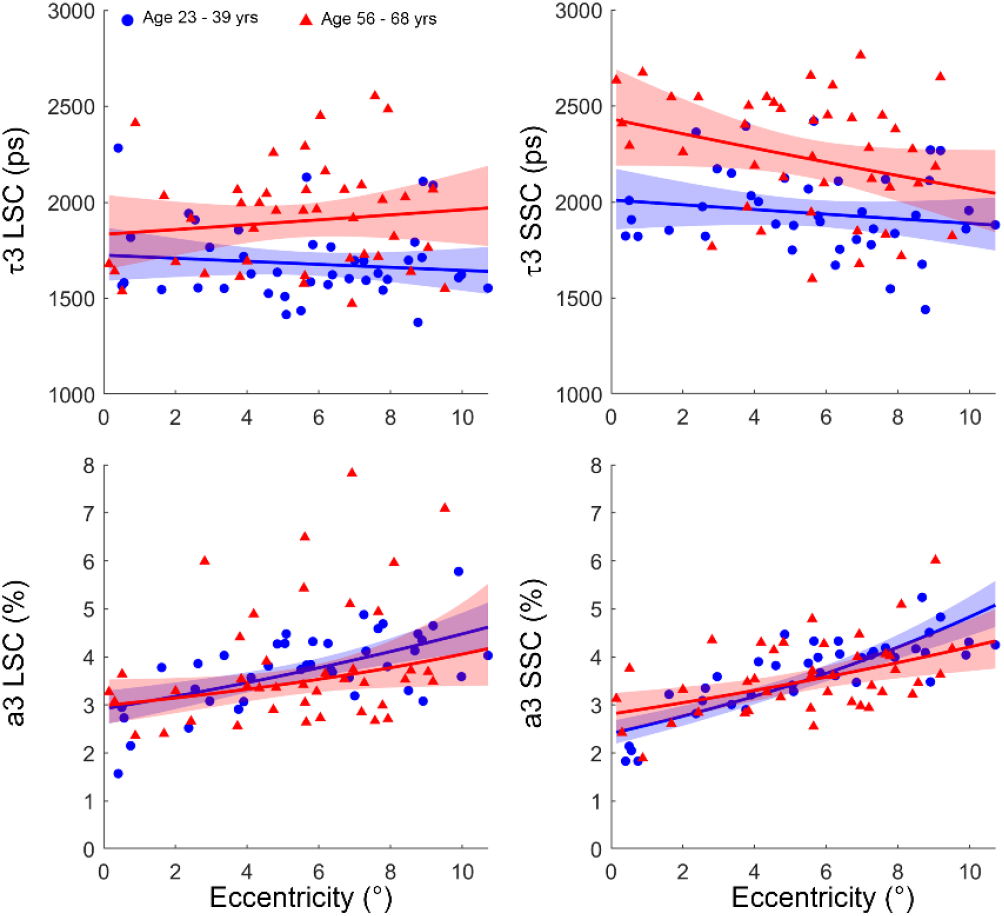
Lens autofluorescence estimated in the long (LSC) and short spectral channel (SSC) across different ages and retinal eccentricities using the S3-exp model.

With respect to retinal eccentricity, *τ*_3_ showed no significant spatial dependence in the LSC (p = 0.141, **Table 3**), whereas a significant decrease was observed in the SSC (p = 0.014, **Table 4, Fig. 4** top panels). By contrast, *a*_3_ exhibited a strong positive association on retinal eccentricity in both channels (all p < 0.001, **Tables 5** and **6, Fig. 4** bottom panels).

### 3.4 AO confocal imaging reduced the weight of lens autofluorescence

When estimated using the S3-exp model, the amplitudes of lens autofluorescence (*a*_3_) were 3.8 ± 1.1% in the LSC and 3.6 ± 0.8% in the SSC. In contrast, evaluation with the 3-exp model yielded *a*_3_ values of 3.1 ± 1.1% in the LSC and 2.7 ± 0.8% in the SSC. Despite the small weight of the third component to the mean fluorescence lifetime, it accounted for 19.4% to 24.1% of the total photons for the 3-exp model (**Table 7**), comparable to the values measured through pseudophakic eyes without autofluorescent lenses using the clinical FLIO instrument.^12,23^

Averaged across all eccentricities, the third component, i.e., lens autofluorescence, imposed a significant impact on the estimation of the mean retinal autofluorescence lifetime. The mean retinal lifetime (*τ*_*m*_) derived from the 2-exp model was substantially longer than the *τ*_*m*12_ obtained by the 3-exp and S3-exp models, due to the influence of lens autofluorescence (**Fig. 5**). Specifically, in the LSC, *τ*_*m*_ from the 2-exp model (312 ± 44.8 ps) was prolonged by 22.8% compared to 3-exp *τ*_*m*12_ (254.0 ± 42.7 ps, p < 0.0001) and by 30.2% compared to S3-exp *τ*_*m*12_ (239.6 ± 38.7 ps, p < 0.0001). Similarly, in the SSC, the *τ*_*m*_ of the 2-exp model (273.5 ± 44.1 ps) was prolonged by 30.7% compared to 3-exp *τ*_*m*12_ (209.2 ± 39.3 ps, p < 0.0001) and by 42.9% compared to S3-exp *τ*_*m*12_ (191.4 ± 33.1 ps, p < 0.0001). Moreover, when comparing the 3-exp and S3-exp models, the 3-exp model yielded significantly longer lifetimes, as it did not account for the early arrival of lens fluorescence. The *τ*_*m*12_of the 3-exp model was prolonged by 5.7% in the LSC (p < 0.001), and by 8.5% in the SSC (p < 0.001).

**Fig. 5.**
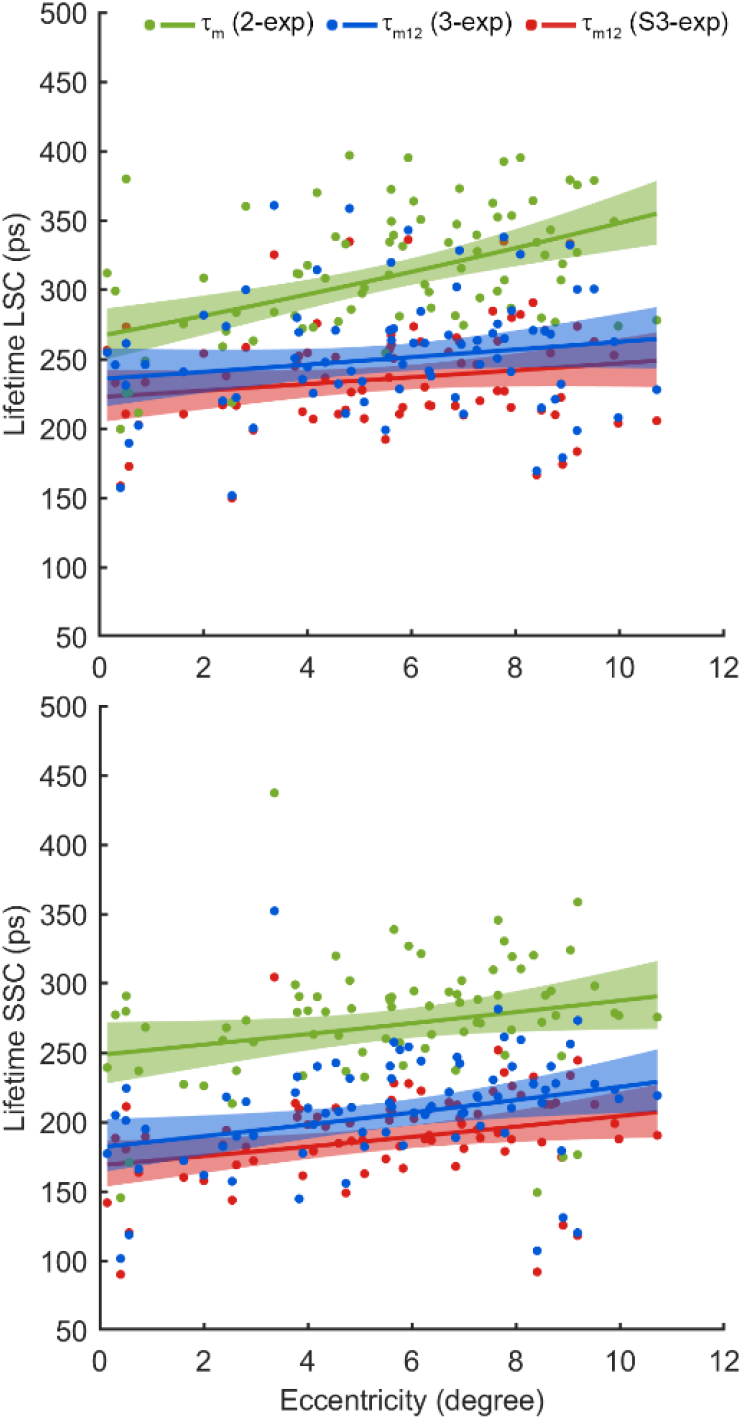
Mean retinal autofluorescence lifetimes estimated using the bi-exponential (2-exp), tri-exponential models with (S3-exp) and without (3-exp) accounting for the early arriving lens signal in the long spectral channel (LSC) and short spectral channel (SSC) across the retinal eccentricities.

## 4. Discussion

In this study, we evaluated the impact of lens autofluorescence on AOFLIO measurements using three mathematical models of fluorescence decay incorporating two- and three-exponential components. We demonstrated that AOFLIO substantially reduced the contribution of crystalline lens autofluorescence; however, residual lens signal remained non-negligible and continued to influence the measured retinal fluorescence lifetime. These findings indicate that minimizing lens autofluorescence is critical for obtaining reliable and accurate retinal fluorescence lifetime measurements.

Our study showed that the conventional 2-exp and 3-exp models produced suboptimal fit at the rising edge of retinal autofluorescence decay curve (**Fig. 1**). This finding agrees with previous FLIO studies, which have shown that lens fluorophores have long fluorescence lifetimes in the nanosecond range.^23,25^ Compared to autofluorescence emitted from the retina-RPE complex, autofluorescence from the lens arrives earlier due to the reduced light path. The S3-exp model, in which the third exponential component is temporally advanced by a time shift, provided optimal and biophysically interpretable modeling of this early component and the overall autofluorescence decay (**Fig. 2**).

Across multiple metrics, the lens effects were greater in the SSC than in the LSC. First, LME analysis revealed that the time shift parameter of lens signal was associated with axial length and age, both are subject-specific factors, in the SSC, whereas no such associations in the LSC (**Tables 1** and **2**). In addition, the SSC exhibited a higher ICC than the LSC (0.48 vs 0.39, **Tables 1** and **2**), indicating greater between-subject variability consistent with heterogeneous lens properties across individuals. Second, fluorescence lifetime differences between 2-exp and S3-exp models were more pronounced in SSC. These results align with prior FLIO studies reporting that SSC is more sensitive to lens fluorescence due to the emission spectra of major lens fluorophores such as AGEs and 3-hydroxykynurenine glucoside (3-OHKG), which fluoresce predominantly between 500–600 nm.^10,20,22^

With the S3-exp model, we found that the fluorescence lifetime of the third component (*τ*_3_) measured by our AOFLIO increased significantly with age in both SSC and LSC (all p < 0.05, **Tables 3** and **4, Fig. 4** top panels). However, the amplitude (*a*_3_), although varied with retinal eccentricity, did not exhibit significant change with age (**Tables 5** and **6, Fig. 4**, bottom panels). These results are in agreement with previous FLIO measurements in normal aging eyes, where lens fluorescence lifetime increases with age.^10,20,23^ The significant increase in the amplitude of the third component with eccentricity may be related to differences in the angular collection efficiency or localized changes in lens transmission or backscattered light from different fundus locations.^23,25^ Specifically, when AOFLIO acquired images at increased retinal eccentricity, the lens became tilted, resulting in a longer optical path and consequently an increased number of autofluorescence photons.

Although lens autofluorescence was not completely eliminated in AOFLIO, our results demonstrate that AOFLIO markedly suppressed the relative contribution of the lens signal compared with conventional FLIO.^12,23^ Adaptive optics increased the number of autofluorescence photons detected from the retina–RPE complex through the confocal pinhole while enhancing spatial rejection of out-of-focus signals. This effect is reflected in the reduced contribution of the third exponential component, as quantified by the q_3_ parameter. In our dataset, when modeled using the 3-exp approach, the mean and range of the q3 were closely matched those reported in FLIO studies of pseudophakic eyes, in which the natural crystalline lens had been surgically removed.^12,23^ For example, Schweitzer and colleagues reported q_3_ values in pseudophakic eyes of 21.6 ± 2.9 ps in the LSC and 24.7 ± 3.2 ps in the SSC,^23^ with an overall range of 22–46 ps.^12^ In our cohort, we measured comparable values of 19.4 ± 5.0 ps (range: 5.4–30.2 ps) in the LSC and 24.1 ± 4.3 ps (range: 14.9–38.4 ps) in the SSC. By contrast, when modeled using the S3-exp framework, our AOFLIO results (22.7 ± 3.6 ps (range: 14.8–32.2 ps) in the LSC and 29.0 ± 2.7 ps (range: 22.3–37.5 ps) in the SSC) fell outside the ranges typically reported for phakic FLIO measurements, which are 25.6–38.8 ps^26^ (mean: 29.5 ± 9.9 ps) ^20^ in the LSC and 48.7–59.9 ps^26^ (mean: 54.2 ± 10.6 ps) ^20^ in the SSC (Table 7). Taken together, AOFLIO achieved a level of lens autofluorescence suppression comparable to that observed in pseudophakic eyes, in which the artificial intraocular lens is non-fluorescent. The substantial reduction in lens contribution is further supported by the absence of age dependence in the lens autofluorescence weight (a_3_) in both spectral channels (**Tables 5 and 6**).

**Table 6.** Associations between a3 and age and retinal eccentricity (short spectral channel)

| Time shift | $\beta$ | CI- | CI+ | SE | *p* value | VIF |
| --- | --- | --- | --- | --- | --- | --- |
| Age | 0.00 | -0.01 | 0.01 | 0.01 | 0.989 | 1.01 |
| Retinal eccentricity | 0.17 | 0.13 | 0.21 | 0.02 | < 0.001 | 1.01 |
| Marginal R<sup>2</sup> | 0.375 |  |  |  |  |  |
| Conditional R<sup>2</sup> | 0.663 |  |  |  |  |  |
| ICC | 0.461 |  |  |  |  |  |

A small amplitude of the third component can lead to large errors in fluorescence lifetime estimation. The amplitude *a*_3_ of the lens-related component in our dataset was within a range of 3% – 4% in both SSC and LSC, which agrees with previous AOFLIO studies that reported *a*_3_ < 5% in healthy phakic eyes.^28^ However, its contribution to total photon-weighted lifetime is disproportionately large, ranging from 19.4% to 29.0% in our data (**Table 7**) and causing overestimation of the fluorescence lifetime by 22.8–42.9% if uncorrected (**Fig. 5**). These results emphasize that even a small intrusion of the lens signal can substantially bias retinal fluorescence lifetime estimation.

**Table 7.** Photon ratio of the third exponential component in phakic and pseudophakic eyes.

| Studies | $q_3$ (%) (3-exp) | | $q_3$ (%) (S3-exp) | |
| --- | --- | --- | --- | --- |
|  | LSC | SSC | LSC | SSC |
| **Phakic eye (this study)** | **$19.4 \pm 5.0$** | **$24.1 \pm 4.3$** | **$22.7 \pm 3.6$** | **$29.0 \pm 2.7$** |
| **(range)** | **(5.4 ~ 30.2)** | **(14.9 ~ 38.4)** | **(14.8 ~ 32.2)** | **(22.3 ~ 37.5)** |
| Phakic eye<sup>12</sup> | 20 ~ 50 | 26 ~ 74 |  |  |
| Pseudophakic eye<sup>12</sup> | 22 ~ 46 | 22 ~ 46 |  |  |
| Pseudophakic eye<sup>23</sup> | $21.6 \pm 2.9$ | $24.7 \pm 3.2$ | | |
| Cataractous eye<sup>23</sup> | $28.4 \pm 6.0$ | $51.3 \pm 10.3$ | | |
| Cataractous eye<sup>26</sup> | 25.4 ~ 37.3 | 44.9 ~ 56.5 | 25.6 ~ 38.8 | 48.7 ~ 59.9 |
| Cataractous eye<sup>20</sup> | | | $29.5 \pm 9.9$ | $54.2 \pm 10.6$ |

Strengths of our study are AOFLIO acquired from a group of well characterized normal phakic eyes, a novel AOFLIO instrument with robust adaptive optics compensation for human ocular aberrations, a systematic evaluation of the autofluorescence lifetime estimated using multiple mathematic models, and comparative analyses across different age groups and retinal eccentricities. Limitations of this study include a relatively small sample size and the absence of subjects with pseudophakic lenses. Additionally, all measurements were conducted using a fixed, relatively large confocal pinhole, which reduced the system’s ability to reject out-of-focus signals. This reflects an inherent limitation of this imaging modality, constrained by light safety requirements and participant tolerance.

Interpretation of the time-shift parameter remains non-trivial. The mean time shift was 122.0 ps in the LSC and 132.9 ps in the SSC, corresponding to ∼18.3–19.8 mm, consistent with the round-trip distance between the lens and retina.^44^ However, the time shift also showed a significant negative association with imaging location (β = −1.70, p < 0.001) in both spectral channels (**Tables 1 and 2**), which cannot be explained by optical path length alone. A plausible explanation is eccentricity-dependent prolongation of retinal autofluorescence lifetimes, whereby increased contribution of long-lifetime components at peripheral locations reduces the separation between the retinal and lens signals, resulting in a smaller estimated shift. This effect may also reflect measurement limitations related to the ∼10–20 ps temporal resolution of AOFLIO and sensitivity to photon counts and fitting accuracy. Further studies employing refined acquisition strategies and larger samples are warranted.

Despite these limitations, our analysis suggests that AOFLIO has achieved near-pseudophakic suppression of lens signal. Future improvements are possible. Smaller confocal pinholes may further enhance axial sectioning and reject out-of-focus signals. Moreover, more advanced imaging system design with ring-shaped aperture or dual-point scanning may be incorporate to minimize lens fluorescence.^25^ Additionally, subject-specific shift calibration based on ocular biometry (e.g., axial length, lens thickness) could improve model accuracy.

In conclusion, AOFLIO is also affected by the inherent autofluorescence emitted by the natural lens. A computational model with 3 exponential functions and compensating for the time difference of the lens signal is necessary for accurate measurement of the retinal autofluorescence lifetime. Minimizing the influence of lens autofluorescence is crucial in studies aiming to accurately measure the metabolic status of the retina, especially in aging populations and disease conditions where subtle changes in autofluorescence lifetime may serve as early indicators of pathology.

## Acknowledgement

ChatGPT (OpenAI) was used to support grammar checking and stylistic editing of the manuscript. The tool did not contribute to the scientific content, data analysis, or interpretation. All final revisions were reviewed and approved by the authors.

## Declaration

This manuscript describes original work that has not been published or submitted elsewhere. A revised version of this work will be submitted to **Translational Vision Science & Technology (TVST)**.

## Data Availability

All data produced in the present work are contained in the manuscript.

